# The importance of identifying red flags in wrist and hand pain: a scoping review protocol

**DOI:** 10.1101/2023.03.23.23285639

**Authors:** Federico Rossi, Marco Cordella, Elena Lanfranchi, Roberta La Marca, Ruggero Mennea, Daniele Pierazzoli, Matteo Cioeta, Leonardo Pellicciari

## Abstract

**Background:** Musculoskeletal pain can be defined as a consequence of a trauma or as a disorder of varying nature, which can be manifested in bones, muscles, tendinous and ligamentous structures, including those of the wrist and hand. However, such pain can mimic pathologies that need further diagnostic investigations; as a consequence, healthcare practitioners have to be able to detect their signs and symptoms, in order to refer the patient to the most appropriate clinician.

The current literature lacks of reviews that analyze red flags in the field of wrist and hand pain. The reviewers would like to look for any correlations that could be useful in a correct screening process by clinicians.

**Methods:** This scoping review will be performed in accordance with the PRISMA extensions for Scoping Reviews (PRISMA-ScR). Studies will be included if they meet the following inclusion criteria: population characterized by wrist and hand pain with or without comorbidities either at the present or in the past. No limits of age, gender and sport activity will be applied. No study design, publication type, and data restrictions will be applied. MEDLINE, Web of Science, Cochrane Library, CINAHL, Embase, PEDro databases will be searched up to February 2023. Two reviewers will independently screen all title, abstracts and full-text studies for inclusion. A data collection form will be developed by the research team to extract the characteristics of the studies included. A tabular and accompanying narrative summary of the information will be provided.

**Conclusions:** This will be the first scoping review to provide a comprehensive overview of the topic. The results will add meaningful information for future research and clinical practice. The results of this research will be published in a peer-reviewed journal and will be presented at relevant (inter)national scientific events.

## Introduction

Musculoskeletal disorders include a wide range of inflammatory and degenerative conditions affecting the muscles, tendons, ligaments, joints, peripheral nerves, and supporting blood vessels. These include clinical syndromes such as tendon inflammations and related conditions (tenosynovitis, epicondylitis, bursitis etc.), nerve compression disorders (carpal tunnel syndrome, sciatica etc), and osteoarthrosis, as well as less standardized conditions such as myalgia, low back pain and other regional pain syndromes not attributable to known pathology. The most involved body regions are the low back, neck, shoulder, forearm and, least but not last, wrist and hand.^1^

Specifically, upper limb musculoskeletal pain is common in adulthood and can be an important cause of disability, it leads to reduced quality of life, and results in a huge financial burden.

For example, shoulder pain affects roughly 7-34% of people in westernised countries^2-3^ while hand-wrist pain affects 9-23%^4^.

In a population survey of working-aged English adults^5^, 48% reported experiencing upper limb pain in the past year, with 14% describing persistent pain (>6 months). Furthermore, the 2016 guideline shows that hand, wrist and forearm symptoms in workers are one of the five most common causes of reported work-related complaints^6^.

Ryall et al.^7^ found that, of those presenting to primary care or physiotherapy with upper limb pain, 42% reported pain in the distal region (elbow, forearm, wrist and hand), with 48% reporting persistent symptoms and 19% describing unremitting pain a year later.

Among the main recommendations for the evaluation and treatment of patients with acute, subacute or chronic disorders of the hand, wrist or forearm is the exclusion of pathologies that require urgent medical evaluation or that need to be better clinical view.

In fact, musculoskeletal pain can sometimes mimic pathologies that are not the responsibility of the physiotherapist, defined as red flags (RFs).

These are signs and symptoms detected during the medical history and physical examination that could be related to a serious underlying disease, indicating the need for further diagnostic tests before the patient can be appropriately managed by the healthcare professional^8^.

The ability to recognize a serious pathology is, therefore, a key component of the practice of the Physiotherapist to provide the most accurate and optimal therapy to the patient.

In the current literature there are no systematic reviews analyzing RF of the wrist-hand area. In the management of pathologies of the wrist-hand district clinicians can use guidelines^6^, which, however, do not elaborate on the process of screening for differential diagnosis.

According to the recent literature there is a strong need to investigate which are the possible RFs affecting the wrist-hand district to improve the process of differential diagnosis of serious diseases by health professionals.

The objective of this study is, therefore, to identify and evaluate the main musculoskeletal pathologies at the wrist and hand level which, however, may not be the responsibility of the physiotherapist and which, therefore, require a referral to other health professionals.

## Methods

This study will be prepared using the Joanna Briggs Institute guidelines^10-11^ and following the Preferred Reporting Items for Systematic Reviews and Meta-analysis Protocol extension for Scoping Reviews (PRISMA-ScR)^12^.

### Research question

We formulated the following research question: “What are the non-muscoloskeletal pathologies that mimic musculoskeletal disorders in the wrist and hand?”

### Inclusion criteria

Studies will be considered eligible for inclusion if they meet the Population, Concept and Context (PCC) criteria.

### Population

This scoping review will consider studies that include a population presenting with hand-wrist pain, attributable to red flag, in the presence and absence of present or past comorbidities, regardless of age and gender, sports athletes and non-athletes.

### Concept

This scoping review will consider studies examining different pathologies that mimic musculoskeletal disorders in the wrist-hand region.

### Context

This scoping review will consider studies conducted in any setting.

### Sources

Primary studies with any type of design will be included. No time, geographical or study setting limitations will apply. No language restrictions will apply.

### Exclusion criteria

Studies involving patients with shoulder or elbow-related symptoms will be excluded.

### Search strategies

The search will be conducted across 7 databases (Pubmed, Web of Science, Cochrane Library, PEDro database, CINAHL, Embase and Scopus). In addition, a manual search will be carried out via cross-referencing, on the bibliographies of other relevant reviews and other sources of gray literature (Google Scholar). Authors whose documents are missing useful information for this piece of research will be contacted. The complete search strategy will be reported according to PRISMA-S^13^ (extension for literature searches in systematic reviews) and will be detailed an additional file.

### Selection of studies

The selection process will consist of two levels of screening using the Rayyan QCRI^14^ online software: a title and abstract and (2) a full-text screening. For both levels, two reviewers (RM and DP) will independently screen the articles to determine if they meet the inclusion/exclusion criteria. In case of disagreements, these will be resolved by a third reviewer (FR). The reasons for the exclusion will be recorded and reflected in the supplementary materials. The entire selection process will be reported and presented in a PRISMA flowchart^15^.

### Data extraction

The data extraction process will be carried out by two reviewers (RM, DP) independently using a standardized data extraction table, in which the following have been considered: title and author, objective, typology and result of the study. In case of disagreement between the two reviewers, the third reviewer (FR) will be consulted for a final decision. (Table 3)

**TABLE 1:**
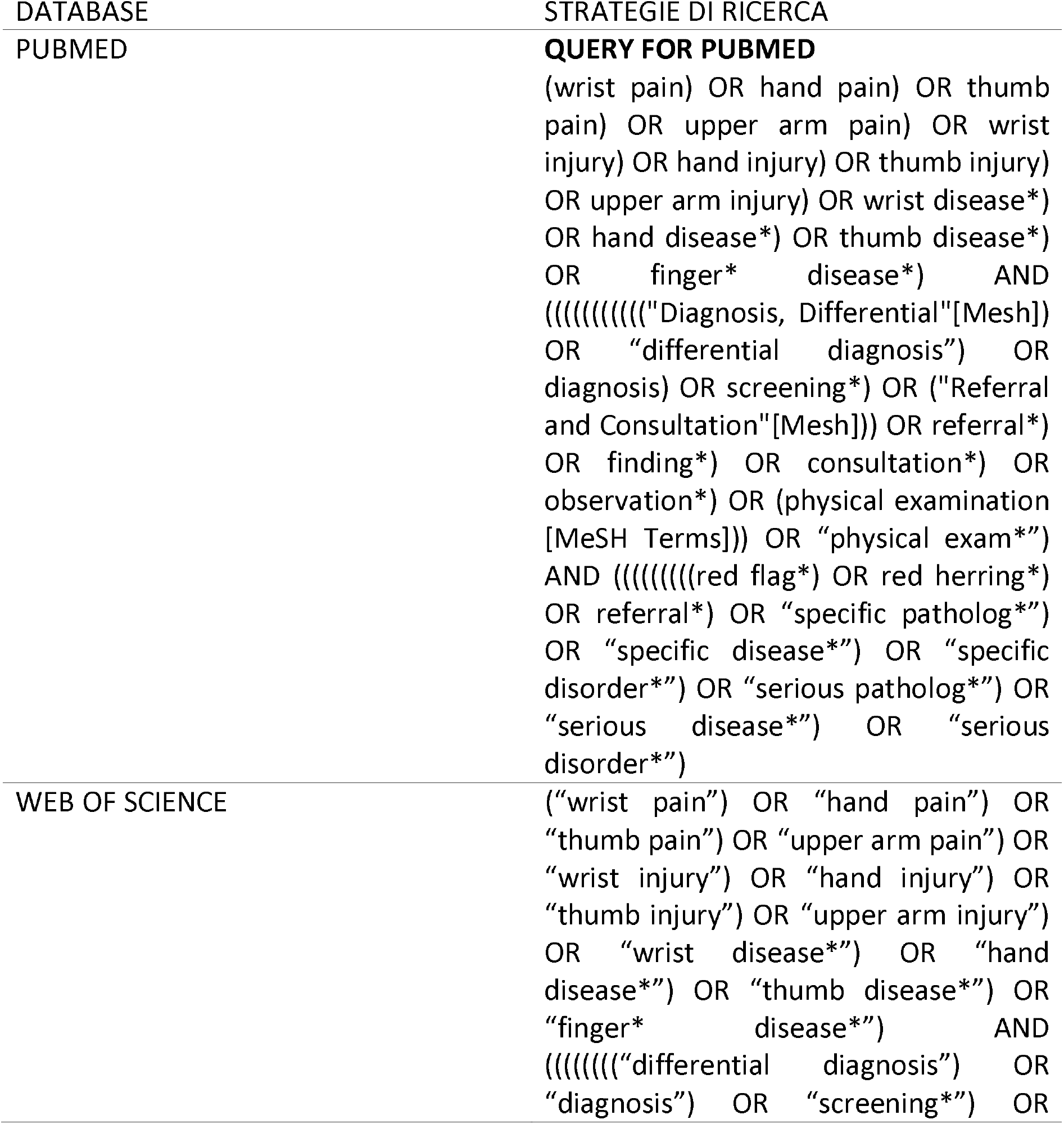
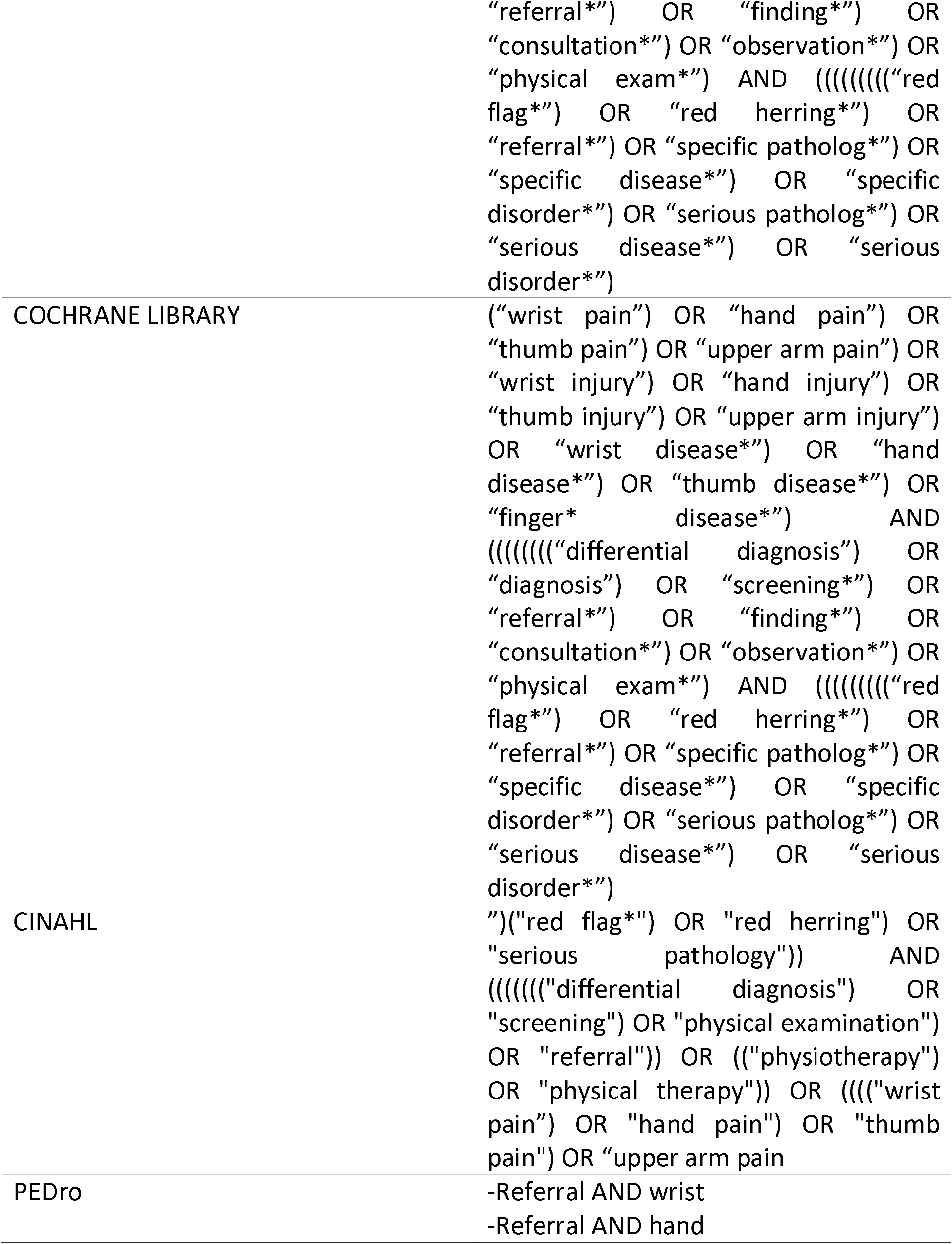
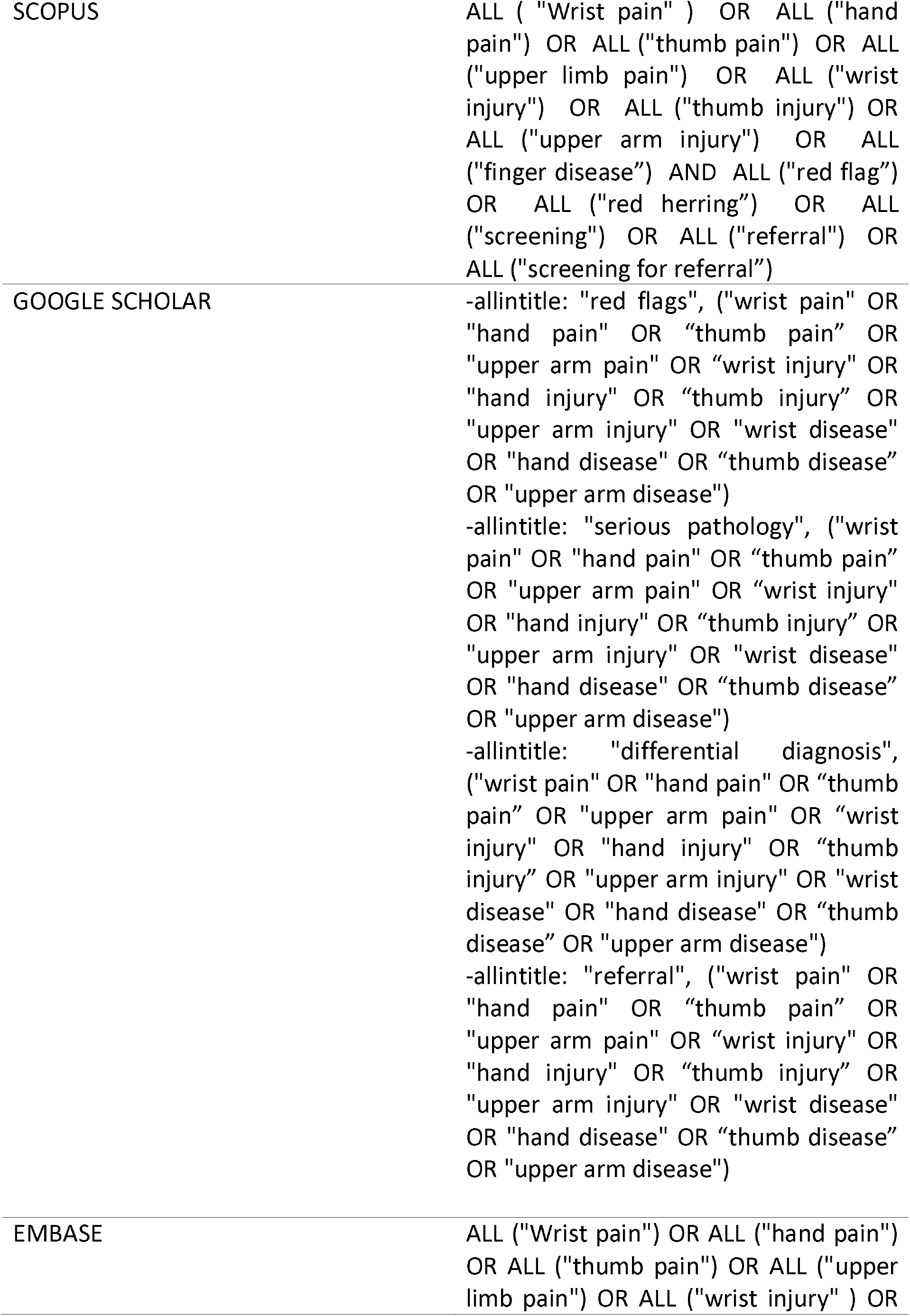
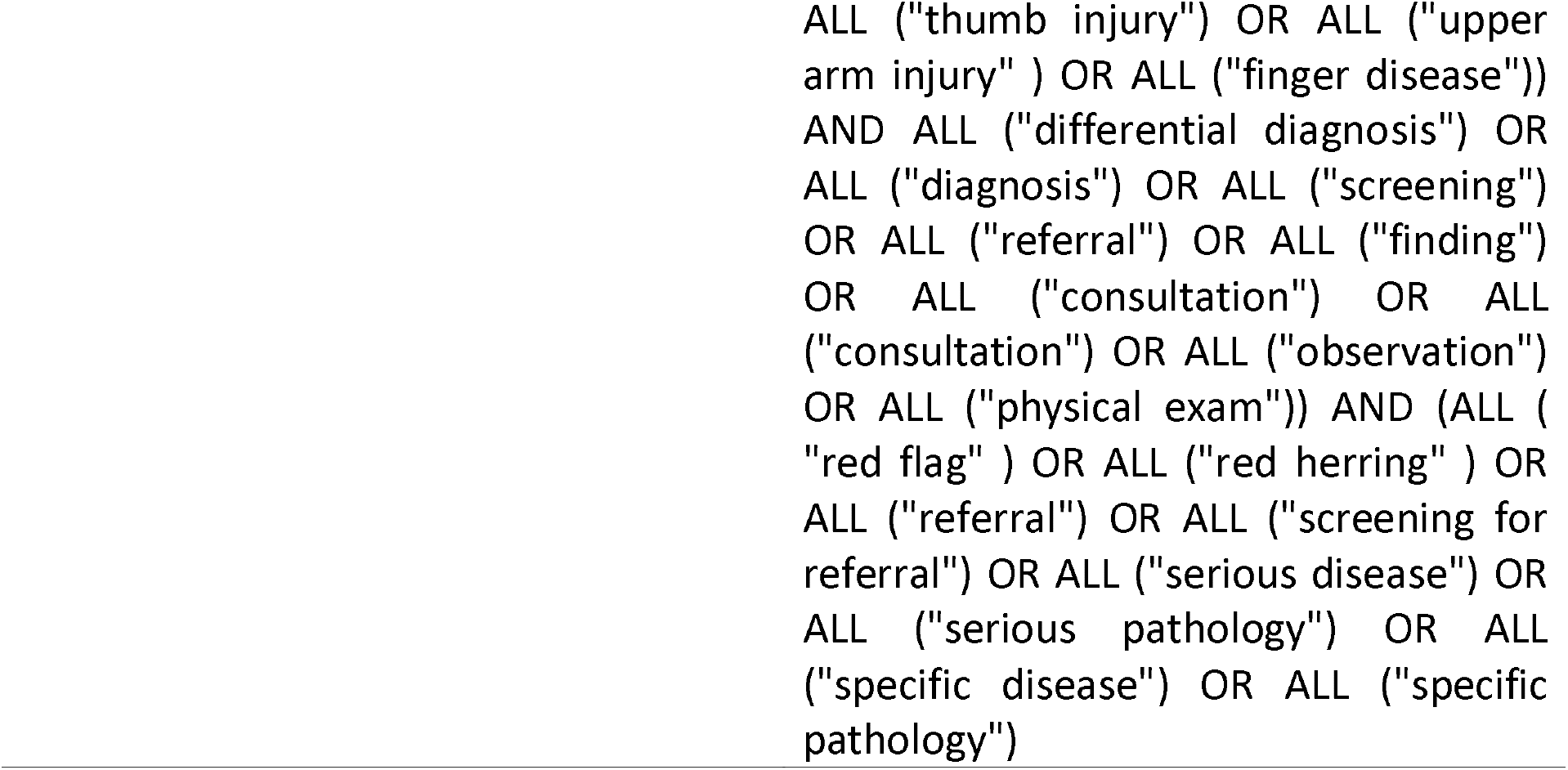
research strategies for each database.

**TABLE 2:** Population, Concept and Context (PCC)) of the scoping review.

|  | INCLUSION CRITERIA | EXCLUSION CRITERIA |
| --- | --- | --- |
| **Population** | Population presenting with hand-wrist pain in the presence and absence of present or past comorbidities, regardless of age and gender, sports athletes and non-athletes. | patients with symptoms related to the shoulder or elbow. |
| **Concept** | studies examining various pathologies that mimic musculoskeletal disorders in the wrist-hand area. | - |
| **Context** | studies conducted in any setting. | - |

**TABLE 3:**
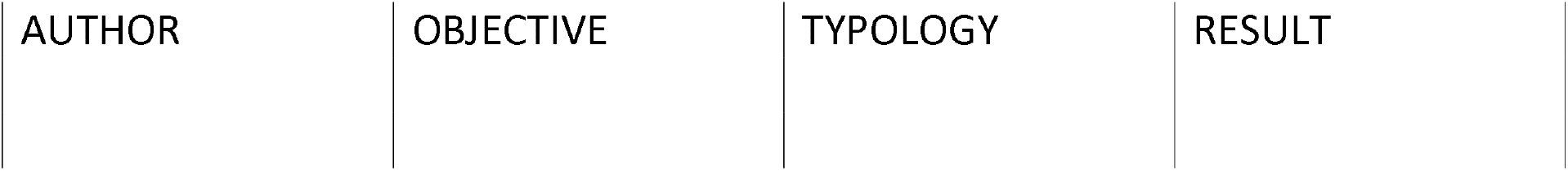
Data extracted from the included studies

### Data management

As a scoping review, the purpose of this study is to aggregate the findings and present an overview on the research rather than to evaluate the quality of the single studies. The results will be presented in two ways:

1. Numerically. Data extraction will be summarized in tabular form. Further categories may be added if considered appropriate.
2. Thematically. A descriptive analysis will be performed pertaining to themes and key concepts relevant to the research questions and according to subgroups that could emerge.

## Data Availability

All data produced in the present work are contained in the manuscript

## Declarations

### Ethics approval and consent to participate

Not applicable.

### Consent for publication

Not applicable.

### Availability of data and materials

Not applicable.

### Authors’ contributions

All authors conceived, designed, drafted and approved the final manuscript.

## Acknowledgements

Not applicable.

## Ethics and dissemination

A manuscript with results will be prepared and submitted for journal publication upon project completation. The findings of the study will be disseminated at a relevant (inter)national conference. The results of this research will be published in a relevant peer-reviewed journal in the rehabilitation and physical therapy field. All results of this scoping review will also be announced at (inter)national scientific events in the area of rehabilitation of musculoskeletal disorders.

## List of abbreviations

RFs: Red Flags
PCC: Population-Concept-Context
JBI: Joanna Briggs Institute
PRISMA: Preferred Reporting Items for Systematic Reviews and Meta-Analyses

